# Language impairment in autistic adolescents and young adults: Variability by definition

**DOI:** 10.1101/2025.09.05.25335184

**Authors:** Teresa Girolamo, Lindsay Butler, Julia Parish-Morris

## Abstract

**Purpose:** Co-occurring language impairment (LI) in autism is common and predicts long-term academic, occupational, and social outcomes. Yet little is known about LI in autism beyond childhood. One challenge to closing this gap is the lack of consensus regarding how LI should be operationally defined in adolescents and adults. This study examines how different epidemiological definitions of LI influence clinical classification and observed language profiles in speaking autistic adolescents and young adults.

**Method:** Participants (*N* = 75; ages 13-30) varying in levels of autism traits completed norm-referenced measures of overall expressive language, overall receptive language, receptive vocabulary, expressive vocabulary, nonword repetition, and nonverbal intelligence. Scores were compared to epidemiological definitions for LI varying in criteria and cutoffs from-1 *SD* to-1.5 *SD*. Data were analyzed using descriptives and clustering.

**Results:** More stringent definitions yielded a greater proportion of participants meeting LI criteria, and more stringent cutoffs for LI yielded greater overall consistency in clinical classification across individual language measures, but there was no one-to-one ratio between cutoff and clinical classification. Clustering indicated three profiles differentiated on the basis of language and nonverbal cognitive skills, but each cluster was heterogeneous. Individual performance also varied across language measures.

**Discussion:** Findings support multi-domain approaches to characterizing language skills in autistic adolescents and adults, including those with LI. Future work is needed to understand language skills in autism beyond childhood and how to develop effective assessment practices.

## Language impairment in autistic adolescents and young adults: Variability by definition

There is growing evidence of heterogeneity in language abilities across the autism spectrum (Bal et al., 2016; Butler et al., 2023; Pizzano et al., 2024), but little is known about language beyond childhood (Howlin & Taylor, 2015). Language skills are associated with long-term academic, occupational, and social outcomes (Brignell et al., 2018; Magiati et al., 2014), underscoring the importance of accurately characterizing language skills across development. In the United States, the transition to adulthood is a key developmental stage, as autistic youth age out of child-and school-based services (“Individuals with Disabilities Education Improvement Act [IDEIA] of 2004,” 2018). Many lack access to services and supports (Eilenberg et al., 2019; Roux et al., 2024; Weir et al., 2022). Despite these challenges, language in autistic adolescents and young adults remains poorly understood.

More than 50% of autistic individuals have structural language impairment (Boucher, 2012), and over 60% show broader language delays (Baird et al., 2006; Levy et al., 2010).

However, operational definitions of language impairment (LI) vary substantially (Girolamo et al., 2023), limiting comparisons across studies and developmental stages. In this report, LI refers to difficulties in one or more domains of structural language (e.g., morphosyntax, semantics, phonology) (Schaeffer et al., 2023), and is treated as a construct versus a diagnostic category.

This distinction is important, as existing classification systems provide limited operational guidance for identifying LI in older individuals. The *Diagnostic and Statistical Manual of Mental Disorders, 5^th^ Ed.* (DSM-5) includes LI as a specifier but does not define measurement criteria or clarify how LI should be operationalized (American Psychiatric Association [APA], 2013). Similarly, the *International Classification of Diseases, 11^th^ Revision* (ICD-11) includes severity qualifiers for functional language impairment without specifying assessment procedures (World Health Organization [WHO], 2022).

As a result, the literature uses varying assessment batteries and cutoff thresholds (Girolamo et al., 2023; Koegel et al., 2020; Kwok et al., 2015; Magiati et al., 2014), often relying on too little information, such as using verbal IQ as an indicator of overall cognitive ability (McCauley et al., 2020), which confounds language with cognition (Grondhuis et al., 2018), or using a label like “LI” as an indicator of language skills across domains (Schaeffer et al., 2023). These practices complicate the interpretation and comparison of findings across studies and influence how language abilities are documented. A secondary implication of lack of operational guidance is that after publication of the DSM-5 (APA, 2013), students with a primary disability label of autism may not receive speech-language evaluation or access to services (Musgrove, 2015), even though U.S. legislation mandates access to services in all areas associated with a disability and their unique needs (“IDEIA of 2004,” 2018).

The present study examines how different epidemiological definitions of LI influence classification outcomes and observed language profiles in speaking autistic adolescents and young adults. By applying multiple empirical definitions to the same sample, a focus is on how measurement decisions influence classification and interpretation. Consistent with recent calls for multidimensional approaches to assessment (Kover & Abbeduto, 2023; Schaeffer et al., 2023), this study evaluates language performance across multiple language domains and examines how classification differs across definitions. To better understand heterogeneity within this population, this study also explores patterns of language and nonverbal intelligence.

### Structural LI in Autistic Adolescents and Adults

Characterizing LI in autism beyond childhood presents both conceptual and methodological challenges. While there are validated clinical language assessments for youth (Girolamo et al., 2022; Nitido & Plante, 2020), far fewer measures are normed for adults over age 21 (Manenti et al., 2023). Understanding performance on norm-referenced assessments is important, as they are often used to document language skills in service systems (Burke et al., 2024; Selin et al., 2019). In practice, studies often use assessments designed for other clinical populations. These practices introduce uncertainty regarding interpretation of receptive and expressive language profiles, as operational definitions of LI use specific cutoffs.

Limitations of available adult language measures further complicate interpretation. Some assessments were normed decades ago on non-representative samples, such as adults with a college education (Hammill et al., 2007), limiting the generalizability of norms. Receptive and expressive vocabulary tests normed for younger populations have yielded age equivalent scores outside basal and ceiling ranges when administered to autistic adults ranging widely in age (ages 21-64 years), leaving it unclear how to interpret assessment results (Howlin et al., 2004, 2014; Mawhood et al., 2000). At the same time, reliance on a single language domain, such as vocabulary, may underestimate broader structural language difficulties, independent of autism (Calder et al., 2023; Schaeffer et al., 2023). Elsewhere, omnibus (i.e., overall receptive-expressive) language batteries developed for adults with acquired language disorders (e.g., aphasia) have been applied to autistic adults without brain injury (Lewis et al., 2008), raising questions about construct validity. These examples illustrate the limitations of available tools for measuring LI as a multidimensional construct.

A second challenge involves variability in how LI is operationalized in adolescent and adults. Studies have used differing combinations of measures and cutoffs, often extending norms to those over age 21 years. Definitions have ranged from-2 *SD* on omnibus language composite (Botting, 2020), to-1.5 *SD* on omnibus language composite (Clegg et al., 2021; Norbury et al., 2016), to-1 *SD* on receptive vocabulary or omnibus language composite scores, or-2 *SD* on ≥ 2 subtests of an omnibus language measure (Johnson et al., 1999b; Poll et al., 2010). In autistic samples, LI has been also defined using-1.25 *SD* on two or more measures spanning overall receptive language, overall expressive language, vocabulary, and phonological domains (Girolamo & Rice, 2022), consistent with epidemiological approaches developed in nonautistic youth (Tomblin et al., 1996). Across studies, there is no consensus on which measures should be included, which cutoffs are most appropriate, or how norms should be applied in adulthood.

Characteristics of prior adult samples are summarized in Supplementary Table 1.

Independent validation studies also demonstrate that there is no single operational definition for LI in adulthood. In the Ottawa longitudinal study, differences between test norming samples and the study cohort motivated the development of local norms to define LI, inclusive of autism (Johnson et al., 1999b). A-1 *SD* cutoff on receptive vocabulary or omnibus language yielded higher LI estimates (12.6%) than expert clinical judgment (8.3%) (Johnson et al., 1999a), emphasizing how classification thresholds can meaningfully alter prevalence estimates. In a separate study, multi-domain combinations of phonological, semantic, and syntactic measures yielded the highest classification accuracy for developmental LI in nonautistic adults (Fidler et al., 2011, 2013). In contrast, single measures of narration, nonword repetition, sentence production, and grammatical judgment were inconsistent in identifying LI (Fidler et al., 2011). While narration can confound structural language skills with autism traits (Baixauli et al., 2016), prior work suggests that LI and autism traits are separable constructs (Loucas et al., 2008; Manenti et al., 2024; Silleresi et al., 2020). Together, findings support use of multi-domain, multi-method assessment.

A related methodological issue concerns the use of intelligence quotient (IQ) cutoffs in studies of LI in autism. Many studies exclude individuals below a specified IQ threshold (Girolamo et al., 2023), despite current diagnostic frameworks allowing co-occurring autism and intellectual disability (APA, 2013; WHO, 2022). The use of IQ exclusion criteria may constrain observed patterns of LI and limit representation of the full autism spectrum (Russell et al., 2019; Shaw et al., 2025). To this end, recent work in autistic adults demonstrated no one-to-one correspondence between structural language and nonverbal IQ (NVIQ), with heterogeneous profiles indicating individuals with LI and average of high NVIQ (≥110), as well as individuals with lower NVIQ (<80) and without LI (Manenti et al., 2024). Importantly, LI in that study was defined using cutoff thresholds (i.e., moderate at-1.25 *SD* and severe at-1.65 *SD*) on non-normed nonword repetition and sentence repetition measures relative to a comparison sample of non-autistic adults (Manenti et al., 2024), illustrating how operational decisions influence observed patterns. Overall, variability in LI cutoffs, inclusion criteria, and measurement choices motivates a dimension approach to characterizing language in autistic adolescents and adults.

### The Current Study

In the absence of consensus of how to operationalize LI in adolescence and adulthood, examining how classification varies by definition can clarify the influence of measurement decisions. This exploratory, observational study applies established epidemiological definitions of LI to the same sample of speaking autistic adolescents and young adults. By varying only the operational criteria in terms of measures included and cutoff thresholds, this approach serves to isolate how different definitions shape classification outcomes and interpretation. Consistent with prior epidemiological work (Johnson et al., 1999a; Norbury et al., 2016; Tomblin, 1996), this study evaluates LI using normed measures across multiple language domains, including omnibus expressive and receptive language, vocabulary, and phonology. Following current diagnostic frameworks that recognize variability across domains (APA, 2013; WHO, 2022), this study did not use NVIQ exclusion criteria. Research questions were:

1) To what extent does clinical classification of LI differ by definition in speaking autistic adolescents and young adults: (a)-1 *SD* on omnibus language or receptive vocabulary measures, or-2 *SD* on ≥2 subtests of an omnibus language measure (Johnson et al., 1999a); (b)-1.25 *SD* on ≥2 measures of overall receptive language, overall expressive language, receptive vocabulary, expressive vocabulary, and nonword repetition, or on an omnibus language measure (Tomblin et al., 1996); and c)-1.5 *SD* on ≥2 measures from (b) (Norbury et al., 2016), and by individual language measure?
2) How does performance on language measures and NVIQ relate within this population? Based on prior validation studies indicating that stricter cutoffs yield greater concordance with clinical judgment (Johnson et al., 1999a), it was expected that more stringent definitions would yield lower LI estimates. Given evidence that vocabulary may be a relative linguistic strength in autism and may not fully capture structural language difficulties (Arunachalam & Luyster, 2016; Schaeffer et al., 2023), it was expected that fewer participants would meet LI cutoffs on receptive and expressive vocabulary relative to broader language measures. Finally, consistent with recent work showing dissociation between language and NVIQ in autistic adults (Manenti et al., 2024), it was expected that participants would show heterogeneous profiles.

## Methods

### Participants

Participants were speaking autistic adolescents and young adults aged 13 to 30 years.

This age range was selected to capture the transition to adulthood period, including adolescents approaching transition planning and young adults beyond eligibility for school-based services (“Every Student Succeeds Act,” 2015; “IDEIA of 2004,” 2018). Inclusion criteria were: (a) meeting DSM-5 criteria for autism spectrum disorder (APA, 2013), confirmed through a clinical best estimate (CBE) approach (Bishop & Lord, 2023); (b) age between 13 and 30 years; (c) monolingual English speaker, as study activities were in English; (d) adequate hearing and vision for completing audiovisual assessment tasks; and (e) primary use of spoken language to communicate, as activities required verbal responses. Autism diagnosis was confirmed using a CBE procedure that triangulated documentation of a prior professional autism diagnosis, Social Responsiveness Scale-2 (SRS-2) (Constantino & Grubler, 2012) overall *t*-scores, and expert clinical judgment by a trained clinician. SRS-2 caregiver and self-report forms were combined, because mean overall T-scores did not significantly differ across form types (*p* =.061): caregiver student forms (*n* = 37), caregiver adult forms (*n* = 15), and adult self-report forms (*n* = 21).

### Procedures

This study received institutional review board approval from San Diego State University (HS-2023-0225-SMT) and recruited participants using a multi-step process. The team shared study flyers with organizations providing services to autistic individuals, provided personalized consultation about the study to potential participants and caregivers, and obtained informed consent and assent prior to data collection. Participants provided informed consent if they were their own legal guardian; otherwise, caregivers provided consent and participants provided assent. Recruitment and data collection took place from 2022 to 2024 remotely on HIPAA-compliant Zoom. Participants completed assessments using the digital version of assessments, in accordance with test publisher telepractice guidance (Pearson, 2025). Examiners followed manualized administration procedures and completed training prior to data collection. Language and NVIQ measures were administered in a standardized order within one or more sessions, depending on participant fatigue and scheduling needs. A trained research assistant independently verified scoring accuracy for all measures, except those automatically scored through publisher platforms (e.g., SRS-2, Raven’s 2).

## Measures

### Demographics

Participants or caregivers reported participant race and ethnicity using U.S. Census categories (Office of Management and Budget, 1997). Respondents could select multiple categories or provide write-in responses. Respondents also reported participant sex assigned at birth and gender, with optional write-in responses.

### Autism Traits

Autism traits were assessed using SRS-2 (Constantino & Grubler, 2012) caregiver or self-report forms as appropriate for age. The SRS-2 consists of 65 items rated on a four-point Likert scale and yields standardized T-scores (*M* = 50, *SD* = 10), reflecting social communication and interaction and restricted interests and repetitive behaviors. Internal consistency for forms ranges from α =.95 to.97. T-scores are categorized as subclinical (59), mild (60-65), moderate (66-75), and severe (≥76). Per study procedures, SRS-2 scores were used as part of the CBE process for confirming autism status and were also analyzed dimensionally.

### Language Skills

Language was assessed across multiple domains using norm-referenced, untimed, direct behavioral assessments. Omnibus language (a composite measure derived from multiple subtests) was assessed using six subtests of the Clinical Evaluation of Language Fundamentals–Fifth Edition (CELF-5) (Wiig et al., 2013). The CELF-5 provides scaled scores (*M* = 10, *SD* = 3) and composite indices (*M* = 100, *SD* = 15) for expressive, receptive, and overall expressive-receptive language (composite reliability *r* =.95-.96). Expressive subtests included: Formulated Sentences (sentence production; *r* =.86), Recalling Sentences (sentence repetition; *r* =.94), and Semantic Relationships (sentence-level semantic processing; *r* =.89). Receptive subtests included: Word Classes (semantic associations; *r*:.90), Understanding Spoken Paragraphs (discourse-level comprehension; *r* =.85), and Sentence Assembly (sentence-level integration; *r* =.93). For participants over age 21 (*n* = 24), age 21 norms were used, consistent with prior work (Botting, 2020; Clegg et al., 2021; Fidler et al., 2011).

Receptive vocabulary was assessed using the Peabody Picture Vocabulary Test–Fifth Edition (Dunn, 2019), and expressive vocabulary using the Expressive Vocabulary Test–Third Edition (EVT-3) (Williams, 2019). These co-normed measures provide standard sores (*M* = 100, *SD* = 15; reliability *r* =.97) and are normed from early childhood through >90 years. The PPVT-5 requires participants to select one of four images corresponding to a spoken word. The EVT-3 requires participants to generate a single-word response to an auditory question and visual stimulus.

Phonological processing was assessed using the Syllable Repetition Task (SRT) (Shriberg et al., 2009a). Participants repeat 18 nonwords of two to four syllables, which use early-acquired phonemes to minimize articulation and speech motor confounds (/b/, /d/, /m/, /n/, and /a/) (Shriberg et al., 2009a). Percent accuracy was used as the primary outcome variable.

Reliability for 16-year-olds, the oldest norming group, ranges from *r* =.69 to.89 (*M* = 93.2, *SD* = 3.0) (Shriberg et al., 2009b; Shriberg & Mabie, 2017).

### NVIQ

NVIQ was assessed using the Raven’s Progressive Matrices–Second Edition (Raven et al., 2018), long form (*M* = 100, *SD* = 15; reliability *r* =.88-.91). The Raven’s is untimed and minimizes verbal demands by requiring participants to identify the missing element in a visual matrix without any spoken instructions. This measure reduces confounding of language ability with cognitive assessment (Grondhuis et al., 2018).

### Data Processing and Analysis

All analyses were conducted using SPSS 31.0 (IBM Corp., 2025) using an α level of.05. Because several variables violated assumptions of normality, nonparametric measures were used.

### Data Preparation

A trained research assistant independently checked scoring for all measures except those automatically scored by publisher platforms (e.g., SRS-2, Raven’s 2). Next, data were examined for missingness. As missing data were minimal (<5%), autism trait scores were replaced using single imputation with predictive mean matching (Little & Rubin, 2019). One participant missing both language and NVIQ scores was excluded from analyses. Prior to analysis, variables were examined for outliers using standardized *z*-scores (|z| > 3.29), boxplots, and Mahalanobis distance for multivariate analyses. No outliers were identified. All observations were retained to maintain representation of variability within the sample. For analyses examining relationships across measures, language, autism trait, and NVIQ scores were standardized (*z*-scored) to allow comparability across scales (Jolliffe & Cadima, 2016).

### Analyses

To examine whether LI classification differed by definition, participants were classified (yes/no) under each of three epidemiological definitions: (a)-1 *SD* on CELF-5 core language or PPVT-5, or <-2 *SD* on ≥2 CELF-5 core language subtests (Johnson et al., 1999a); (b)-1.25 *SD* on ≥2 measures: CELF-5 Receptive Language Index, CELF-5 Expressive Language Index, PPVT-5, EVT-3, and SRT accuracy (Tomblin et al., 1996); and (c)-1.5 *SD* on ≥2 measures from (b) (Norbury et al., 2016). Cochran’s Q tests (Cochran, 1950) evaluated differences in classification rates. To test if LI classification differed by individual language measure, participants were also classified at-1 *SD*,-1.25 *SD*, or-1.5 *SD* thresholds on each language measure: PPVT-5, EVT-3, SRT accuracy, and CELF-5 subtests (Formulated Sentences, Recalling Sentences, Semantic Relationships, Word Classes, Understanding Spoken Paragraphs, Sentence Assembly). Pairwise comparisons were conducted using Dunn’s (1964) procedure, with a Bonferroni (1936) correction for multiple comparisons.

To explore patterns among language, autism trait, language, and NVIQ, analysis used principal component analysis (PCA) followed by clustering. Given sample size and to avoid selective reporting (Dalmaijer et al., 2022), analyses were exploratory and descriptive (Husson et al., 2010). Prior to PCA, assumptions of linearity, sampling adequacy, multicollinearity, and multivariate outliers were evaluated (variance inflation factor ≥ 10). No cases were removed.

PCA was conducted on *z*-scored SRS-2 domains, CELF-5 subtests, PPVT-5, EVT-3, and SRT scores (Abdi & Williams, 2010). Component retention was guided by eigenvalues > 1, scree plot inspection (Cattell, 1966), and interpretability. Varimax rotation was used to aid interpretation.

As autism traits loaded weakly on the primary component, clustering analyses were conducted on language and NVIQ variables only. Agglomerative hierarchical clustering using Ward’s method (1963) and squared Euclidean distance was conducted first to examine cluster structure (Bouguettaya et al., 2015; Day & Edelsbrunner, 1984). Inspection of the dendrogram and agglomerative schedule informed the number of solutions and initial centers entered into *k*-means clustering (Hartigan & Wong, 1979; Lu et al., 2008). *K*-means clustering was conducted with 999 iterations and no running means to ensure stability of cluster assignment. Internal cluster validity was evaluated using the KO_MACROS (Orlov, 2024). External validity was evaluated via discriminant function analysis and Cohen’s κ (1960) comparing *k*-means and hierarchical solutions (Dudoit & Fridlyand, 2002). Patterns were interpreted descriptively based on relative mean *z*-scores across language and NVIQ measures. Effects were interpreted as: small at.25, medium at.55, and large at.95 (Gaeta & Brydges, 2020). Because clustering was exploratory and sample dependent, findings were interpreted as illustrative of heterogeneity versus evidence of discrete profiles. Participants were classified into NVIQ bands based on test developer guidance and clinical criteria indicating that an IQ of 70 to 75 represents clinically significant differences (American Association on Intellectual and Developmental Disabilities, 2024; Raven et al., 2018): low (≤75), borderline (76-89), average (90-109), and high (≥110).

## Results

### Sample Characteristics

The final sample included 75 participants aged 13 to 30 years; see Tables 1 and 2. One participant missing both language and NVIQ scores was excluded from analysis. SRS-2 scores were missing for two participants. Given minimal missingness (Little & Rubin, 2019), scores were imputed using predictive means matching. Participants represented diverse racial and ethnic backgrounds. The sample had a female-to-male ratio for sex at birth and gender of one to 2-2.1 in the overall sample, one to 2.2-2.3 among participants 21 years or younger, and of one to 1.6 among participants over age 21. The mean SRS-2 overall T-score fell within the “moderate” range, indicating clinically elevated autism traits.

**Table 1.**
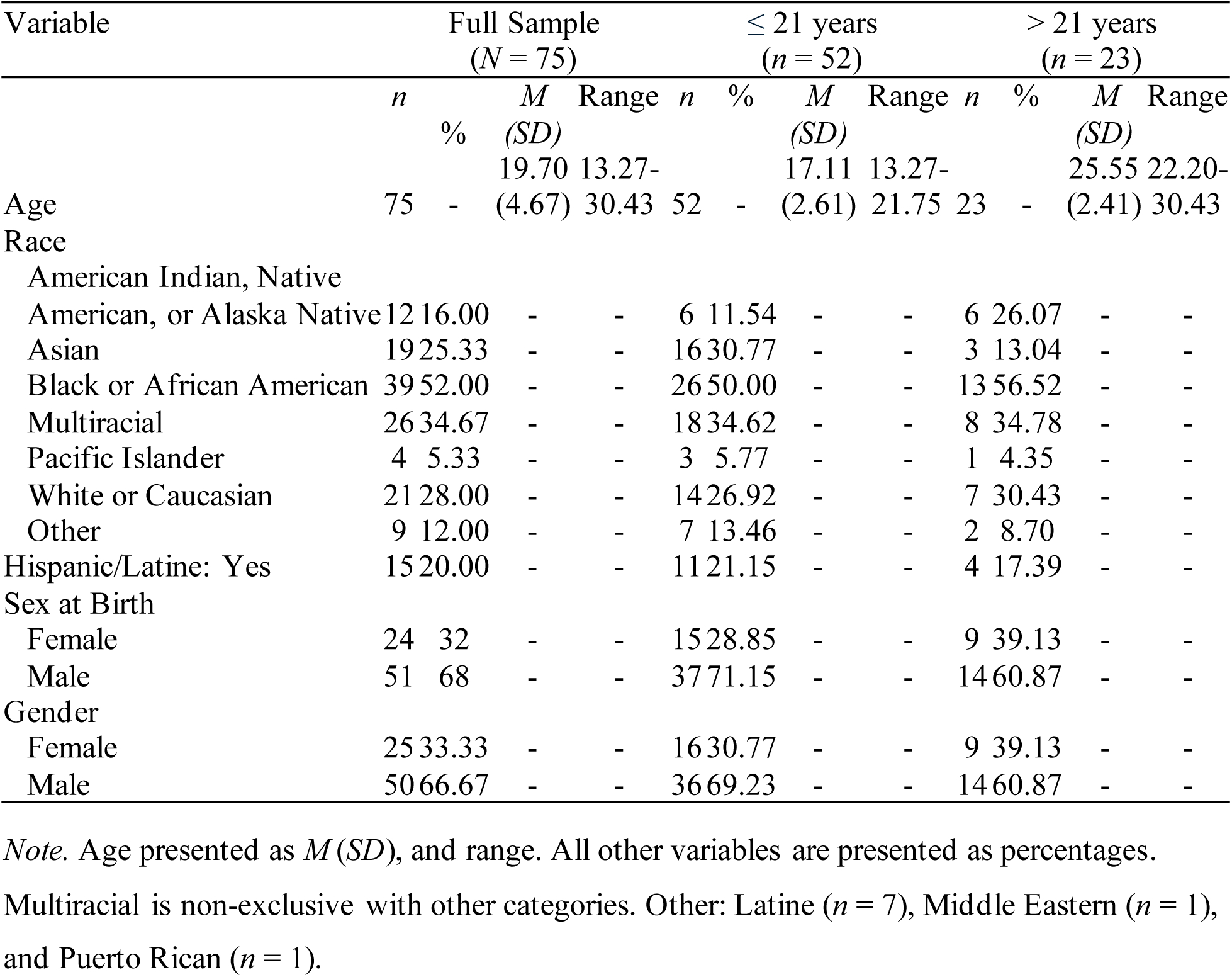
Sample Characteristics (N = 75)

**Table 2.**
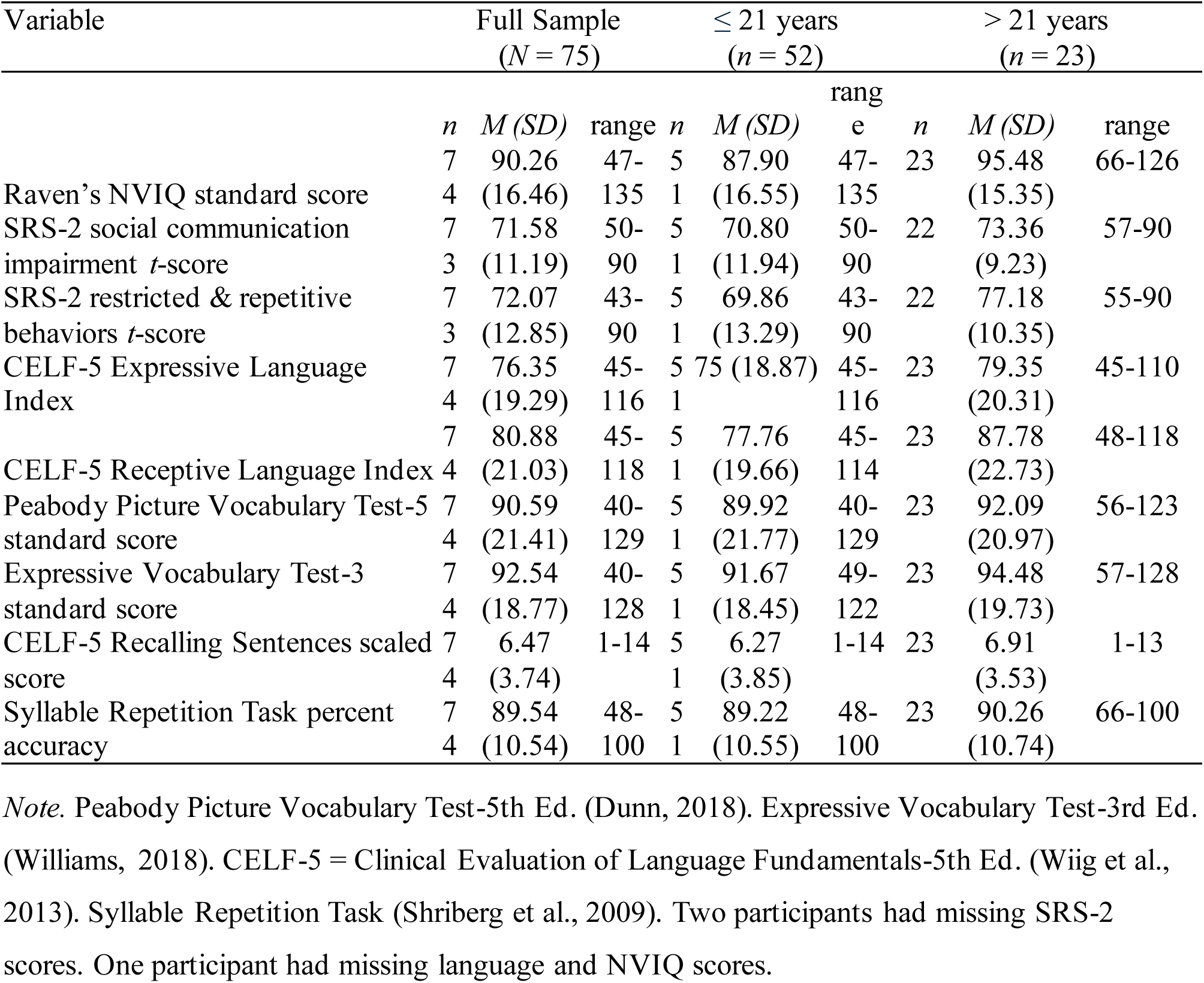
Sample Clinical Assessment Scores (N = 75)

### LI Classification by Definitions and Measure

Clinical classification of LI differed significantly by definition, χ^2^(2)=19.54, *p* <.0001.

Classification rates were: (a) 70.27% using the-1 *SD* on CELF-5 core language score or PPVT-5 scores, or-2 *SD* on ≥2 CELF-5 core language subtests (Johnson et al., 1999a); (b) 62.16% using the-1.25 *SD* on ≥2 measures (CELF-5 Receptive Language Index, CELF-5 Expressive Language Index, PPVT-5, EVT-3, and SRT overall accuracy) (Tomblin et al., 1996); and (c) 52.70% using the-1.5 *SD* on ≥2 measures from definition (b) (Norbury et al., 2016). Yet only the decrease from (a) to (c) was significant (17.57%), *p* <.0001. Thus, greater cutoff stringency was associated with lower LI classification, but differences were not uniformly proportional across cutoff thresholds.

More stringent cutoffs generally yielded greater consistency in LI classification across language measures, as indicated by fewer significant pairwise comparisons:-1 *SD* (*n* = 18),-1.25 *SD* (*n* = 13), and-1.5 *SD* (*n* = 9); see Supplementary Table 2 and Supplementary Figure 1.

However, classification varied by measure; see Figure 1. CELF-5 Understanding Spoken Paragraphs yielded higher LI classification rates at less stringent thresholds (-1 *SD* and-1.25 *SD*) relative to several other measures, such as EVT-3 (Δ: 33.78-40.54%), PPVT-5 (Δ: 25.68-35.13%), CELF-5 Sentence Assembly (Δ: 18.92-29.73%), and CELF-5 Word Classes (Δ: 21.62-32.43%). In contrast, EVT-3 yielded comparatively lower LI classification rates across cutoffs, including CELF-5 Sentence Repetition (Δ: 20.27-21.62%). Across cutoffs, no single measure consistently classified participants as meeting or not meeting LI definitional criteria. Findings indicate that LI classification is sensitive to cutoff and to specific language domains assessed.

**Figure 1.**
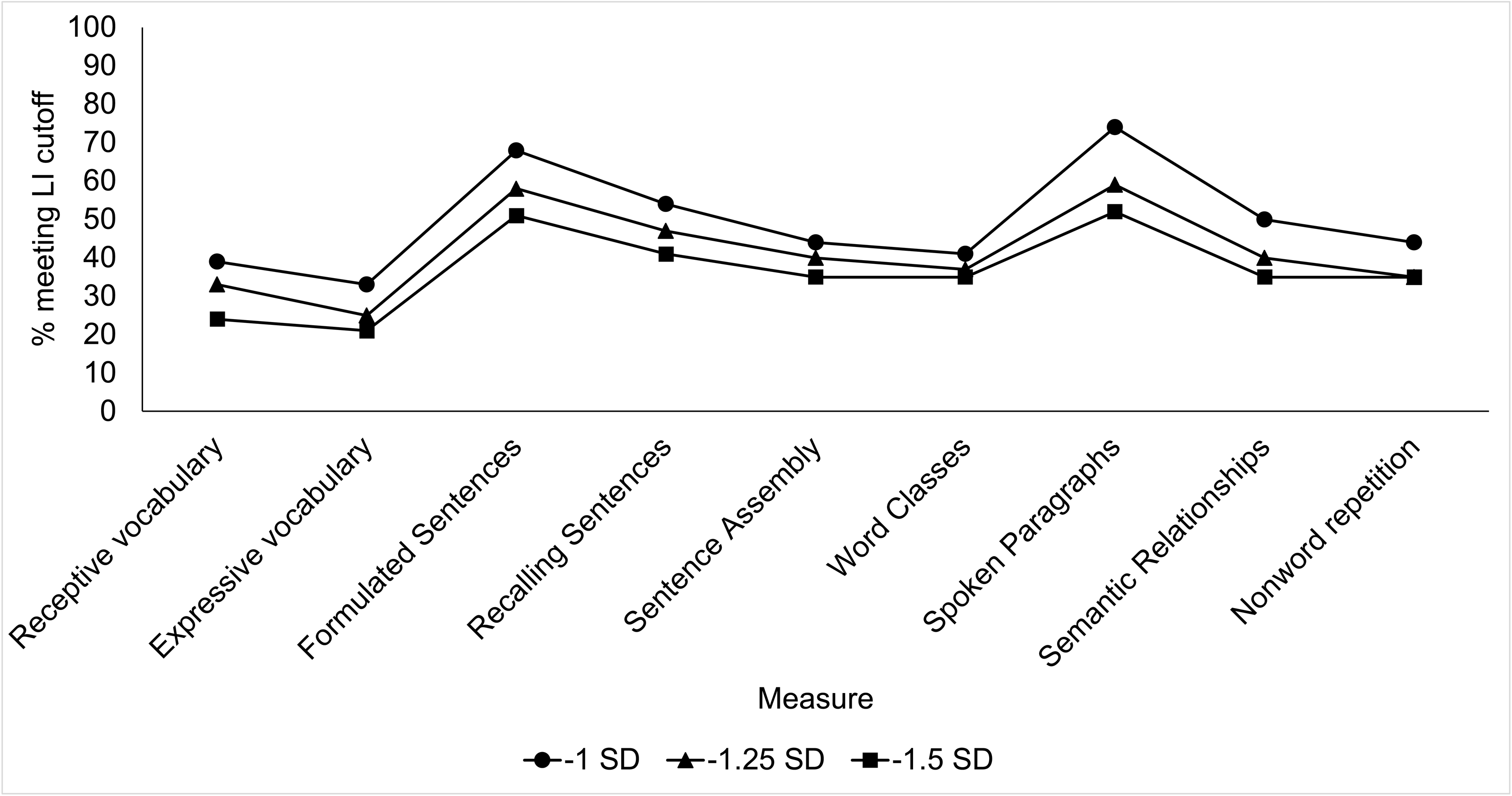
Percent participants meeting language impairment cutoffs across language measures. LI = language impairment. Receptive vocabulary was assessed by the Peabody Picture Vocabulary Test-5 (Dunn, 2019). Expressive vocabulary was assessed by the Expressive Vocabulary Test-3 (Williams, 2019). Formulated Sentences, Recalling Sentences, Sentence Assembly, Word Classes, Understanding Spoken Paragraphs, and Semantic Relationships were assessed by the Clinical Evaluation of Language Fundamentals-Fifth Edition (Wiig et al., 2013). Nonword repetition was assessed by the Syllable Repetition Task overall percent accuracy (Shriberg et al., 2009).

### Language Performance in NVIQ Patterns

PCA assumptions were met. All variables had at least one *r* > 0.3, sampling was adequate (KMO = 0.89) (Kaiser, 1974), and Bartlett’s (1951) test was significant (*p* <.0001). Two components with eigenvalues >1 accounted for 73.28% of the total variance. Following Varimax rotation, language measures and NVIQ loaded strongly on Component 1 (50.32%), while autism traits loaded strongly on Component 2 (24.99%); see Supplementary Table 3 and Supplementary Figure 2. Because autism traits weakly associated with the primary language and NVIQ component (*r* <.16), clustering analyses were run on only language and NVIQ variables.

Hierarchical clustering suggested a three-cluster solution, which was confirmed using *k*-means clustering; see Supplementary Figure 3 and Supplementary Table 4. Internal validity metrics and near-perfect agreement between clustering methods, κ =.873, 95% CI [.775,.971], *p* <.0001, supported selection of the three-cluster solution; see Supplementary Table 5. Discriminant function analysis classified 93.24% of cases currently under cross-validation. Of 14 participants expected to be in Cluster 1, one was in Cluster 3; of 31 expected to be in Cluster 2, one was in Cluster 3, and; of 29 expected to be in Cluster 3, three were in Cluster 2.

Clusters differed descriptively in mean language, NVIQ, and sex at birth distribution; see Table 3. Cluster 1 (*n* = 14) demonstrated the lowest mean language scores (<-2 *SD*) and NVIQ scores (<-1.5 *SD*). Cluster 2 (*n* = 29) had language scores (<-1 *SD*) and NVIQ slightly below average (∼-1 *SD*). Cluster 3 (*n* = 31) had language scores at or above the mean (≥-1 *SD*) and average NVIQ. Despite differences in cluster means, there was substantial within-cluster variability; see Figure 2 and Supplementary Figure 4. When examining clinical categories descriptively, there was overlap between LI and NVIQ levels across clusters. Participants meeting more stringent LI criteria (-1.5 *SD* on ≥2 measures) were distributed across NVIQ bands, including low NVIQ (Cluster 1: *n* = 6, Cluster 2: *n* = 5), borderline NVIQ (Cluster 1: *n* = 5, Cluster 2: *n* = 4, Cluster 3: *n* = 1), and average NVIQ (Cluster 1: *n* = 3, Cluster 2: *n* = 11, Cluster 3: *n* = 3). A small number of participants with high NVIQ also met LI criteria at the-1.25 *SD* on ≥2 measures threshold (Cluster 2: *n* = 3, Cluster 3: *n* = 1). These distributions indicate that classification did not align completely with NVIQ category.

**Figure 2.**
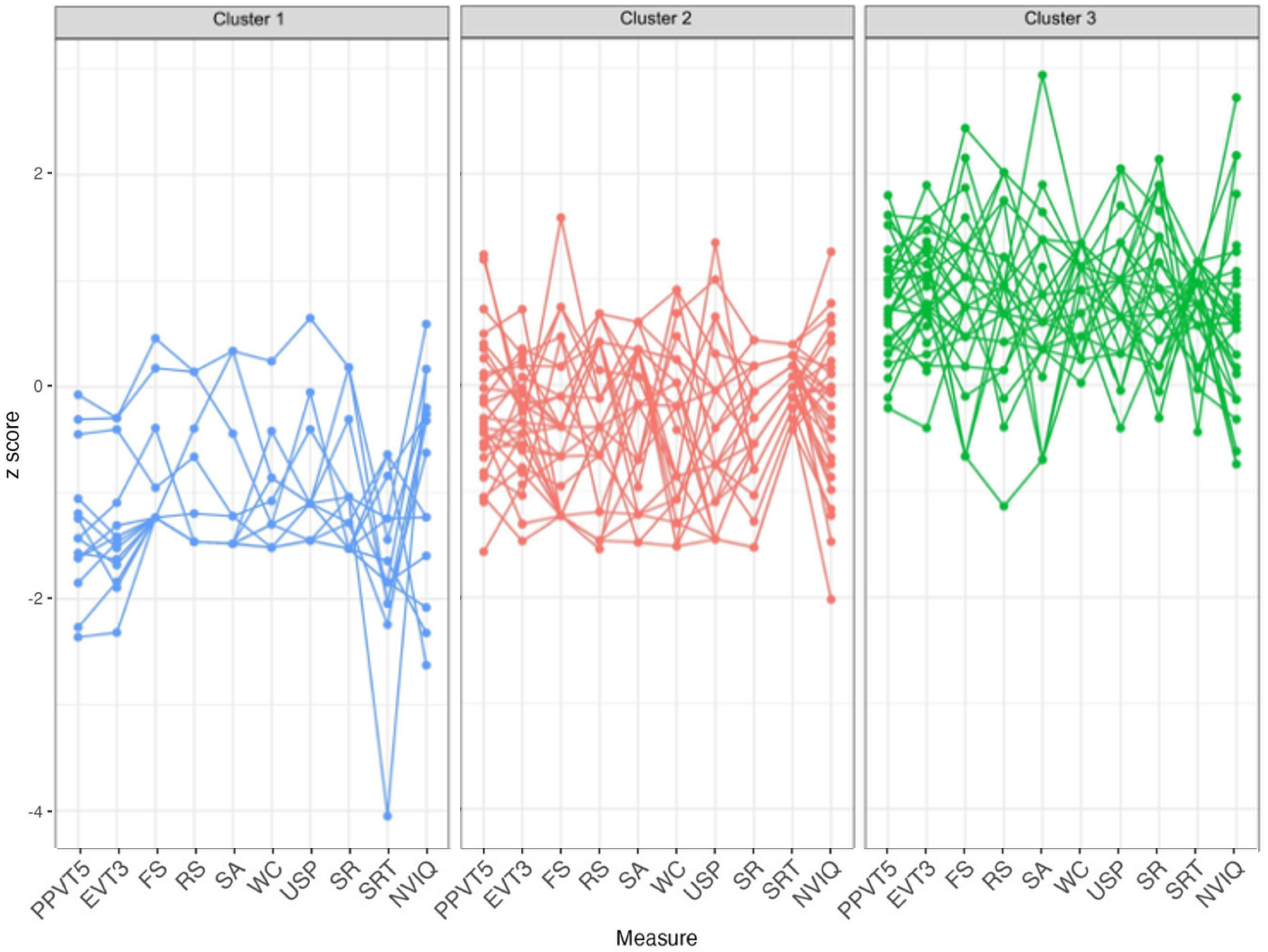
*Z*-scores of individual participants across language and NVIQ measures by cluster. PPVT-5 = Peabody Picture Vocabulary Test-5 (Dunn, 2019). EVT-3 = Expressive Vocabulary Test-3 (Williams, 2019). FS = Formulated Sentences, RS = Recalling Sentences, SA = Sentence Assembly, WC = Word Classes, USP = Understanding Spoken Paragraphs, and SR = Semantic Relationships from the Clinical Evaluation of Language Fundamentals-5 (Wiig et al., 2013). SRT = Syllable Repetition Task (Shriberg et al., 2009). NVIQ = Raven’s 2 nonverbal intelligence quotient (Raven et al., 2018).

**Table 3.** K-Means Three-Cluster Solution Cluster Means, Minimum, and Maximum Scores.

| Measure | Cluster 1<br>(*n* = 14) | Cluster 2<br>(*n* = 29) | Cluster 3<br>(*n* = 31) |
| --- | --- | --- | --- |
| Sex at birth ratio (female:male) | 3:11 | 8:21 | 12:19 |
| Age | 20.03 (13.48-29.67) | 19.09 (13.27-29.03) | 20.26 (13.35-30.43) |
| NVIQ standard score | 76.36 (47-100) | 85.97 (57-111) | 100.55 (78-135) |
| Receptive vocabulary standard score | 62.36 (40-89) | 85.48 (57-117) | 108.13 (86-129) |
| Expressive vocabulary standard score | 67.64 (49-87) | 86.24 (65-106) | 109.68 (85-128) |
| Nonword repetition percent accuracy | 75.23 (48-96) | 90.36 (68-100) | 94.52 (80-100) |
| Word Classes scaled score | 2.79 (1-9) | 6.03 (1-12) | 11.97 (8-14) |
| Formulated Sentences scaled score | 2.07 (1-7) | 3.93 (1-11) | 8.23 (3-14) |
| Recalling Sentences scaled score | 2.43 (1-7) | 5.16 (1-9) | 9.52 (2-14) |
| Understanding Spoken Paragraphs scaled<br>score | 2.36 (1-7) | 3.86 (1-9) | 7.61 (4-11) |
| Sentence Assembly scaled score | 2.43 (1-8) | 5.72 (1-9) | 9.55 (4-18) |
| Semantic Relationships scaled score | 2.93 (1-8) | 5.48 (1-9) | 10.87 (6-16) |

*Note.* Sex at birth presented as counts. All other variables presented as means, minimum, and maximum. NVIQ, receptive vocabulary, and expressive vocabulary standard score $M = 100$ , $SD = 15$ . Nonword repetition percent accuracy: $M = 93.2$ , $SD = 3.0$ . Word Classes, Formulated Sentences, Recalling Sentences, Understanding Spoken Paragraphs, Sentence Assembly, and
Semantic Relationships scaled score: $M = 10$ , $SD = 3$ . Participants over 21: $n = 5$ in Cluster 1, $n = 13$ in Cluster 2, and $n = 5$ in Cluster 3.

There was also observed within-cluster variability. In Cluster 1, most participants scored <-1.5 *SD* on all language measures, with 64.29% (*n* = 9) scoring <-1.5 *SD* across all language measures and 21.43% (*n* = 3) scoring <-1.5 *SD* on all but two of nine measures. However, some participants showed more selective profiles, with relative strengths on vocabulary (EVT-3, PPVT-5) or sentence-level tasks (e.g., CELF-5 Formulated Sentences). Similarly, although most Cluster 2 participants met LI criteria at the <-1.5 *SD* on ≥2 measures (*n* = 26, or 89.66%), some met criteria only at the-1.25 *SD* threshold or showed mixed profiles across measures. In Cluster 3, over two-thirds of participants scored within the typical range on all language measures (*n* = 11, or 35.48%) or all but one measure (*n* = 12, or 38.71%). A subset showed relatively lower scores at-1.25 *SD* or-1.5 *SD* on two to three measures. Across clusters, individual performance varied across language measures and domains, indicating heterogeneity even within groups defined by similar mean language and NVIQ levels.

## Discussion

This study examined how different epidemiological definitions of LI influence classification outcomes and observed language profiles in a sample of speaking autistic adolescents and young adults. Findings indicate that LI classification varied depending upon cutoff, measures, and specific language domains assessed. Exploratory multivariate analyses further supported heterogeneity in profiles, including dissociation between structural language performance and NVIQ in a subset of participants.

### LI Classification and Assessment Performance

LI definitions with more stringent criteria yielded lower LI estimates. However, decreases were not proportional across thresholds, and no single language measures consistently identified participants as meeting LI criteria. These findings align with prior validation studies demonstrating that operational thresholds and measure selection influence LI classification in adulthood (Fidler et al., 2011; Johnson et al., 1999a). LI classification rates in the present sample (52.70%-70.27%) were comparable to epidemiological estimates of structural LI (>50%) and overall LI (63%) in autistic youth (ages 8-10 years) (Baird et al., 2006; Levy et al., 2010).

However, they were higher than those in a recent adult sample (ages 18-56 years; 26.32%) (Manenti et al., 2024). Differences may reflect several factors: the younger age range in this sample (13-30 years), use of norm-referenced measures rather than control-based *z*-scores, broader inclusion of language domains beyond sentence repetition and nonword repetition tasks, and use of an NVIQ measure that did not require use of language. Notably, the 70.27% rate under the-1 *SD* definition (Johnson et al., 1999a) aligns with prior work showing that local norms near this threshold may yield higher LI estimates than expert clinical judgment, particularly when impairment is not present across multiple domains (Johnson et al., 1999b).

Similarly, validation studies in non-autistic adults demonstrate that individual measures vary in discriminatory ability and that multi-domain combinations outperform single measures (Fidler et al., 2011), with adult samples not replicating adolescent-based definitions (Tomblin, 2008).

Together, these findings suggest that variation in LI estimates across studies reflects operational decisions rather than just developmental change.

Although more stringent cutoffs reduced LI classification rates in this study, decreases were not uniform across thresholds or measure. Vocabulary measures yielded lower LI classification rates relative to omnibus expressive-receptive language indices and sentence-level tasks, consistent with concerns that single-domain measures may underestimate broader structural language difficulties (Schaeffer et al., 2023). However, domain-level differences were not consistently significant, and their clinical relevance remains unclear. Determining clinical significance requires a “gold standard” definition for LI in adults, which remains elusive (Fidler et al., 2011; Howlin & Taylor, 2015), and corresponding measurement approaches.

Patterns of language and NVIQ further illustrate heterogeneity. Consistent with prior work (Manenti et al., 2024; Silleresi et al., 2020), language performance and NVIQ did not align uniformly. Descriptively, participants showed different patterns from Manenti et al. (2024), where no autistic adults showed high NVIQ and LI. A small number of participants with high NVIQ met LI criteria, whereas others demonstrated strengths across several language domains. Here, two participants with high NVIQ met LI criteria at <-1.25 *SD* and <-1.5 *SD* on 2 or more measures: CELF-5 Formulated Sentences, Recalling Sentences, Semantic Relationships, and Understanding Spoken Paragraphs. While the first three measures are sensitive to LI in nonautistic youth (Calder et al., 2023), their clinical utility could differ for older autistic individuals, as tasks like sentence production require pragmatic skills (Wiig et al., 2013). Overall, these patterns support a dimensional perspective on language in autism and caution against assuming a singular operational definition of LI beyond childhood within the field.

### Implications for Evidence-Based Practice

Findings have implications for practitioners and researchers who interpret language assessment results in autistic adolescents and adults, with implications for building the evidence base (Durkin et al., 2015). While relevant for service eligibility (Selin et al., 2019), performance on norm-referenced measures indicated that classification outcomes were sensitive to operational definitions for LI. No single measure consistently identified LI across definitions. Contrary to expectations, receptive vocabulary and nonword repetition each had two discrepant classification outcomes from other language measures at-1 *SD* and-1.25 *SD*. Sentence repetition also had more discrepant classification outcomes – and the same number of discrepant classification outcomes as expressive vocabulary – at-1 *SD* than at-1.25 *SD* or-1.5 *SD*.

Results caution against relying on a single measure to make inferences about language skills, consistent with best-practice recommendations emphasizing converging evidence (Sackett et al., 1996). Structural language performance may vary independently of autism traits and NVIQ, as observed in this sample and prior work (Manenti et al., 2024; Silleresi et al., 2020). Though such heterogeneity is not new information, it is important to document that it exists. In contexts where assessment informs documentation and eligibility decisions (Musgrove, 2015), awareness of how definitions and measurement choices influence classification may contribute to more precise interpretation of results, and ultimately, access to services. At the same time, standardized assessments capture only one component of language skills and must consider personal goals and desired supports for language and communication (Burke et al., 2024; Cummins et al., 2020). Interpretation of language performance should consider domains assessed, cutoff, and the broader assessment context, especially given the limited availability of adult-normed language measures.

## Limitations

This study encountered several limitations. First, the sample size was modest, and clustering analyses were exploratory. Hence, findings are highly sample dependent and may not replicate (Cox & Sosine, 2023). Although internal and external validity indices supported the three-cluster solution, replication in larger, multi-site samples is needed before making population-level inferences through age-controlled groups or replication cohorts. Second, the CELF-5 was normed only through age 21. While use of age 21 norms in this study aligned with previous work (Botting, 2020; Clegg et al., 2021; Fidler et al., 2011; Poll et al., 2010), the lack of a consensus definition of LI and of adult-normed language measures limit precision in interpreting language performance beyond early adulthood and calculating psychometrics (e.g., sensitivity, specificity). Third, the study included speaking participants with a professional autism diagnosis, which hinders generalizability. Autistic individuals who use minimal spoken language have clinically relevant variability in their language skills (Bal et al., 2016; Butler et al., 2023), but language assessments requiring verbal responses are not designed for all communication profiles. Finally, this study used direct standardized measures, which do not fully capture functional communication across contexts (Barokova et al., 2020; Butler et al., 2022).

Beyond standardized measures, integrating functional and person-centered, naturalistic measures, as well as medical and intervention history, is needed for a more comprehensive understanding of language abilities in adolescence and adulthood (Sackett et al., 1996).

### Future Directions

Future research should prioritize development and validation of language measures normed for autistic adults, including those without a professional diagnosis. Limited availability of adult-appropriate assessments constrains research comparisons and interpretation. Studies comparing direct assessment with clinical judgment, functional communication, and person-centered measures may help clarify LI beyond childhood and how to assess it (Johnson et al., 1999a; Schaeffer et al., 2023). Given evidence that language, NVIQ, and autism traits may vary independently (Manenti et al., 2024), larger and more diverse samples are needed to examine patterns across adulthood. Such sampling would enable robust analytic approaches, including hold-out samples or replication cohorts (Lombardo et al., 2019), would strengthen confidence in classification frameworks and facilitate building consensus toward a definition for LI beyond childhood. Finally, integrating direct clinical assessment with interaction-based measures may clarify how language skills relate to communication experiences and outcomes (Crompton et al., 2020; Nikolaus & Fourtassi, 2023). Such would support precision-driven approaches to assessment and developing supports that better reflect the priorities of autistic individuals across the lifespan.

## Conclusion

In evaluating language assessment across linguistic domains in autistic adolescents and young adults, this study showed that clinical classification of LI differed by operational definition, measure, and cutoff severity. Levels of language and NVIQ dissociated in approximately one-third of the sample. Exploratory analysis indicated heterogeneity in sample performance, even within participants grouped on the basis of language and NVIQ levels.

Overall, findings support use of multi-domain, multi-measure language assessment and a need for adult clinical language measures.

## Data Availability Statement

Data are unavailable due to ethical restrictions. Information on data structure and analysis can be shared upon reasonable request to the corresponding author and IRB approval.

