## Supplementary Information for "Language impairment in autistic adolescents and young adults: Variability by definition"

| **Table 1**  *Approaches to Measuring Language and LI in Adults* | | | | | | | | |
| --- | --- | --- | --- | --- | --- | --- | --- | --- |
| **Author** | **Population** | **Age** | **IQ cutoff** | **IQ measure** | **LI Def.** | **Lang. Measures** | **Scoring** | **Finding** |
| Ballaban-Gil et al. (1996) | 99 DSM-IV autistic disorder | 12-29.5 | Excluded "severe MR" | Est. functional IQ based on test data, academic achievement & ADL | N/A | Caregiver qualitative report | N/A | IQ WNL: most likely to have improved language. Those with "mild to moderate MR" did not improve in comprehension. |
| Botting (2020) | 83 DLD, 86 TD age peers | 24 | Not reported | WASI-3 PIQ | Primary language difficulties at age 7 | CELF-4 UK composite: WC, RS, FS | 21;11 norms | DLD < TD on CELF-4 (70 vs. 100) & PIQ, but both groups had PIQ WNL (99-112). |
| Clegg et al. (2021) | 44 care leavers | 16-24 | None | N/A | CELF-5 CLS ≤ -1.5 *SD* | CELF-5 UK | NR | CELF-5 UK CLS *Mdn* = 67. 27/44 met DLD criteria. |
| Fidler et al. (2011, 2018) | 36 LD, 30 Hist-S/L, 30 Parent-SLI & 96 controls | 18-49 | NVIQ ≥ 70 | TONI-3 NVIQ | N/A | NWR (Kamhi & Catts, 1986), spelling task, GJ task, CASL Grammatical Morpheme subtest, sentence production task, MTT, CELF-4 WD, PPVT-R, speaking rate task | 21;11 norms | MTT, spelling task & CELF-4 WD consistently contributed to classification accuracy & more strongly in combination versus individually. |
| Howlin et al. (2004) | 68 DSM-IV-TR/ICD autistic disorder | 21-48 | childhood NVIQ ≥ 50 | WISC-R, WPPSI, M-P-S, Leiter, Stanford-Binet, or VABS | N/A | BPVS | AE | 7 scored above ceiling & 4 scored below basal AE. BPVS mean AE was 8.26 years: 48% AE < 6 years, 35% AE = 6-15 years, and 16% AE >15 years. |
| Johnson et al. (1999) | 114 childhood S/LI (inc. ASD) & 128 controls | 18-20 | None | WAIS-R PIQ | TOAL-3 SLQ ≤ -1 *SD* or TOAL-3 subtest ≤ -2 *SD* OR PPVT-R ≤ -1 *SD* | PPVT-R; TOAL-3 overall composite | Local norms | Clinical criteria more stringent than local norms for children (LI estimate: 6.7% vs. 12.6%). |
| Lewis et al. (2008) | 17 autism spectrum disorder (13 DSM-IV AS, 2 "HFA", 1 ASD, 1 autism) & 13 peers without a disability | 18-67 | "no other co-morbid condition" | TONI-2 NVIQ | N/A | WAB; RHLB subtests | Test norms | ASD = TD on NVIQ (91 vs. 99). ASD < TD on WAB and RHLB. Language, but not NVIQ, differentiated ASD subgroups. |
| Mawhood et al. (2000) | 19 autism & 20 receptive DLD who had attended hospital special units or special schools | 21-28 | childhood NVIQ ≥ 70 | WAIS-R PIQ, Raven's Matrices | N/A | BPVS, EOWPVT | AE | Autism PIQ = 82.78 & NVIQ = 86.10. BPVS *M* = 53.2 (37.8). 13 had <10 year AE. On the EOWPVT, ~50% did not reach basal AE (8 years); 10 had AE <10 years. |
| Poll et al. (2010) | 13 SLI & 18 TD peers | 18-25 | PIQ ≥ 80 | WAIS-III PIQ | TOAL-3 SLQ ≤-1 *SD* or 2 SLQ subtests ≤ -2 *SD* OR PPVT-R ≤ -1 *SD* | NWR Task (Dollaghan & Campbell, 1998), CELF-3 RS (item 7 onward), GJ task (Rice & Wexler, 1996) | N/A | NWR, GJ & CELF-3 RS contributed to LI classification. Composite best predicted LI status. |
| *Note.* Language in table directly taken from papers. “MR” = “mental retardation.” ADL = activities of daily living. WNL = within normal limits. DLD = developmental language disorder. TD = typically developing. WASI = Wechsler Abbreviated Scale of Intelligence, Third Ed. (Wechsler, 1999). PIQ = Performance intelligence quotient. CELF-4 UK = Clinical Evaluation of Language Fundamentals-4th Ed. UK (Semel et al., 2006). CLS = core language score. WC = Word Classes. RS = Recalling Sentences. FS = Formulated Sentences. LD = learning disability. Hist-S/L = history of speech/language services. Parent-SLI = parent of child diagnosed with specific language impairment. TONI-2/3 = Test of Nonverbal Intelligence, Third Edition (Brown et al., 1990, 1997). CASL-3 = Comprehensive Assessment of Spoken Language (Carrow-Woolfolk, 1999). MTT = Modified Token Task. PPVT-R = Peabody Picture Vocabulary Test-Revised (Dunn & Dunn, 1981). CELF-4 (Semel et al., 2006). WD = Word Definitions. WISC-R = Wechsler Intelligence Test for Children (Wechsler, 1974). WPPSI = Wechsler Pre-School and Primary Scale of Intelligence (Wechsler, 1990). Merrill Palmer (Stutsman, 1948). Leiter Performance Scales (Levine, 1982). Stanford-Binet (Terman & Merrill, 1961). VABS = Vineland Adaptive Behavior Scales (Sparrow et al., 1984). WAIS-R = Wechsler Intelligence Scale for Adults-Revised (Wechsler, 1981). Raven’s = Raven's Matrices (Raven, 1956). British Picture Vocabulary Scale (Dunn et al., 1982). AE = age equivalent. S/LI = speech or language impairments. Test of Adolescent and Adult Language-3rd Ed. (Hammill et al., 1994). SLQ = spoken language quotient. AS = Asperger syndrome. HFA = "high functioning autism." ASD = autism spectrum disorder. WAB = Western Aphasia Battery (Kertesz, 1982). RHLB = The Right Hemisphere Language Battery (Bryan, 1989). Expressive One Word Picture Vocabulary Test (Gardner, 1982). TOWRE-2 = Test of Word Reading Efficiency-2nd Ed. (Torgesen et al., 2012). Wechsler Adult Intelligence Scales-3rd. Ed. (Wechsler, 1999). | | | | | | | | |

| **Supplementary Table 2**  *Dunn’s Post-Hoc Test Adjusted Significance Levels of Pairwise Comparisons Across Language Measures at -1 SD (Upper), -1.25 SD (Middle), and -1.5 SD (Lower)* | | | | | | | | | |
| --- | --- | --- | --- | --- | --- | --- | --- | --- | --- |
| Variable | 1 | 2 | 3 | 4 | 5 | 6 | 7 | 8 | 9 |
| 1. Peabody Picture Vocabulary Test-5 | - |  |  |  |  |  |  |  |  |
| 2. Expressive Vocabulary Test-3 | 1  1  1 | - |  |  |  |  |  |  |  |
| 3. CELF-5 Formulated Sentences | **<.0001**  **.001**  **<.0001** | **<.0001**  **<.0001**  **<.0001** | - |  |  |  |  |  |  |
| 4. CELF-5 Recalling Sentences | .636  .750  .077 | **.044**  **.008**  **.014** | .636  1  1 | - |  |  |  |  |  |
| 5. CELF-5 Sentence Assembly | 1  1  1 | 1  .397  .657 | **.004**  .096  .166 | 1  1  1 | - |  |  |  |  |
| 6. CELF-5 Word Classes | 1  1  1 | 1  1  .657 | **.001**  **.019**  .166 | 1  1  1 | 1  1 | - |  |  |  |
| 7. CELF-5 Understanding  Spoken Paragraphs | **<.0001**  **<.0001**  **<.0001** | **<.0001**  **<.0001**  **<.0001** | 1  1  1 | **.044**  1  1 | **<.0001**  **.044**  .077 | **<.0001**  **.008**  .077 | - |  |  |
| 8. CELF-5 Semantic Relationships | 1  1  1 | .348  .397  .657 | .091  .096  .166 | 1  1  1 | 1  1  1 | 1  1  1 | **.004**  **.044**  .077 | - |  |
| 9. Syllable Repetition Task | 1  1  1 | 1  1  .657 | **.004**  **.003**  .166 | 1  1  1 | 1  1  1 | 1  1  1 | **<.0001**  **.001**  .077 | 1  1  1 | - |
| *Note.* Adjusted *p*-value used a Bonferroni correction for multiple comparisons. Peabody Picture Vocabulary Test-5th Ed. (Dunn, 2018). Expressive Vocabulary Test-3rd Ed. (Williams, 2018). CELF-5 = Clinical Evaluation of Language Fundamentals-5th Ed. (Wiig et al., 2013). Syllable Repetition Task (Shriberg et al., 2009). | | | | | | | | | |

**Supplementary Figure 1.** Significant pairwise comparisons across language measures by cutoff of -1 *SD*, -1.25 *SD*, or -1.5 *SD*
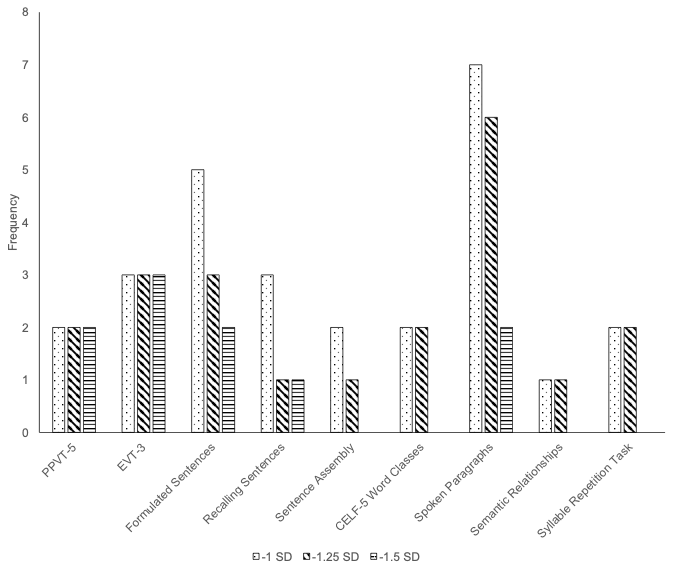

| **Supplementary Table 3**  *Rotated Structure Matrix for Principal Components Analysis of a Two Component Solution* | | | |
| --- | --- | --- | --- |
| Item | Component 1  (50.32%) | Component 2 (24.99%) | Communalities |
| EVT-3 | **0.929** | 0.054 | 0.867 |
| CELF-5 Semantic Relationships | **0.911** | 0.065 | 0.834 |
| CELF-5 Word Classes | **0.874** | 0.100 | 0.774 |
| CELF-5 Recalling Sentences | **0.866** | 0.108 | 0.761 |
| PPVT-5 | **0.863** | 0.082 | 0.751 |
| CELF-5 Understanding Spoken Paragraphs | **0.857** | 0.029 | 0.735 |
| CELF-5 Sentence Assembly | **0.823** | -0.060 | 0.681 |
| CELF-5 Formulated Sentences | **0.794** | 0.173 | 0.661 |
| NVIQ | **0.703** | 0.028 | 0.495 |
| SRT percent accuracy | **0.660** | -0.009 | 0.436 |
| SRS-2 social communication impairment | -0.033 | **0.950** | 0.904 |
| SRS-2 restricted interests and repetitive behavior | 0.153 | **0.934** | 0.895 |
| *Note.* Varimax rotation with Kaiser normalization used. Rotation converged in 3 iterations. EVT-3 = Expressive Vocabulary Test-3rd Ed. (Williams, 2018). PPVT-5 = Peabody Picture Vocabulary Test-5th Ed. (Dunn, 2018). CELF-5 = Clinical Evaluation of Language Fundamentals-5th Ed. (Wiig et al., 2013). SRT = Syllable Repetition Task (Shriberg et al., 2009). SRS-2 = Social Responsiveness Scale-2nd Ed. (Constantino & Grubler, 2012). | | | |

**Supplementary Figure 2.** Scree plot and component plot in rotated space of a principal components analysis run on autism trait, language, and NVIQ z-scores. SCI = Social Responsiveness Scale-2 (SRS-2) social communication & social interaction, RRB = SRS-2 restricted & repetitive behaviors (Constantino, 2012). EVT3 = Expressive Vocabulary Test-3 (Williams, 2018). NVIQ = Raven’s 2 nonverbal intelligence (Raven, 201 8). PPVT5 = Peabody Picture Vocabulary Test (Dunn, 2018). SRT = Syllable Repetition Task (Shriberg et al., 2009). FS = Clinical Evaluation of Language Fundamentals-5 (CELF-5) Formulated Sentences, RS = CELF-5 Recalling Sentences, CELF-5 SA = Sentence Assembly, CELF-5 SR = Semantic Relationships, CELF-5 USP = Understanding Spoken Paragraphs, WC = CELF-5 Word Classes (Wiig et al., 2013).

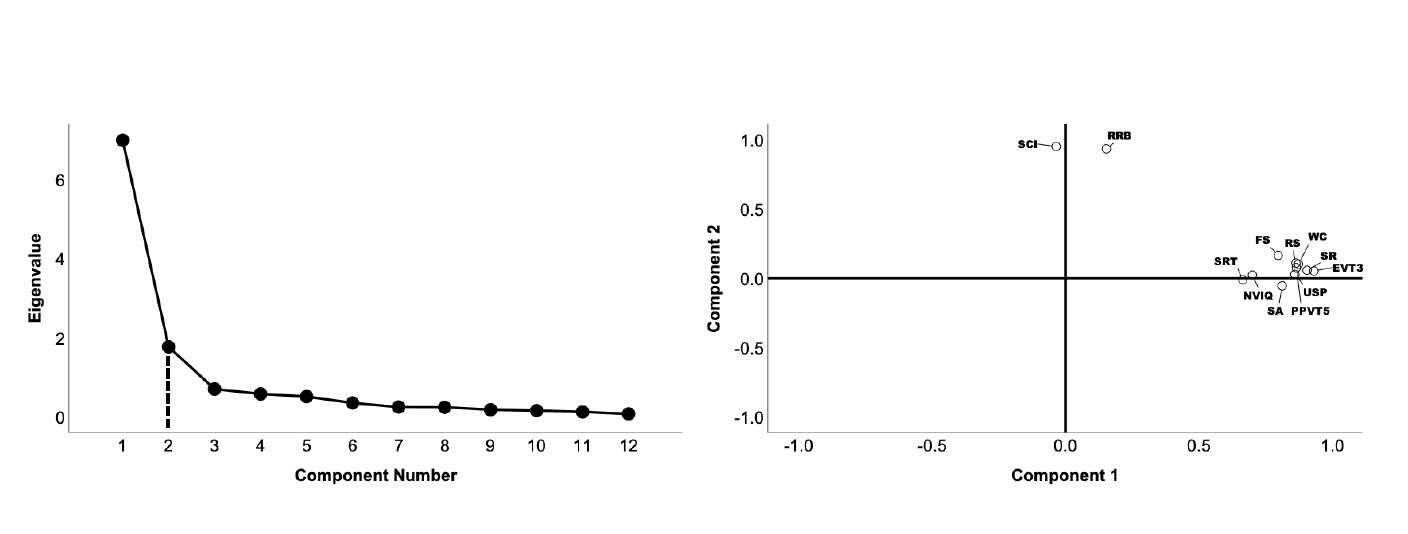

**Supplementary Figure 3.** Dendrogram with line suggesting stopping location and scree plot for agglomerative hierarchical clustering

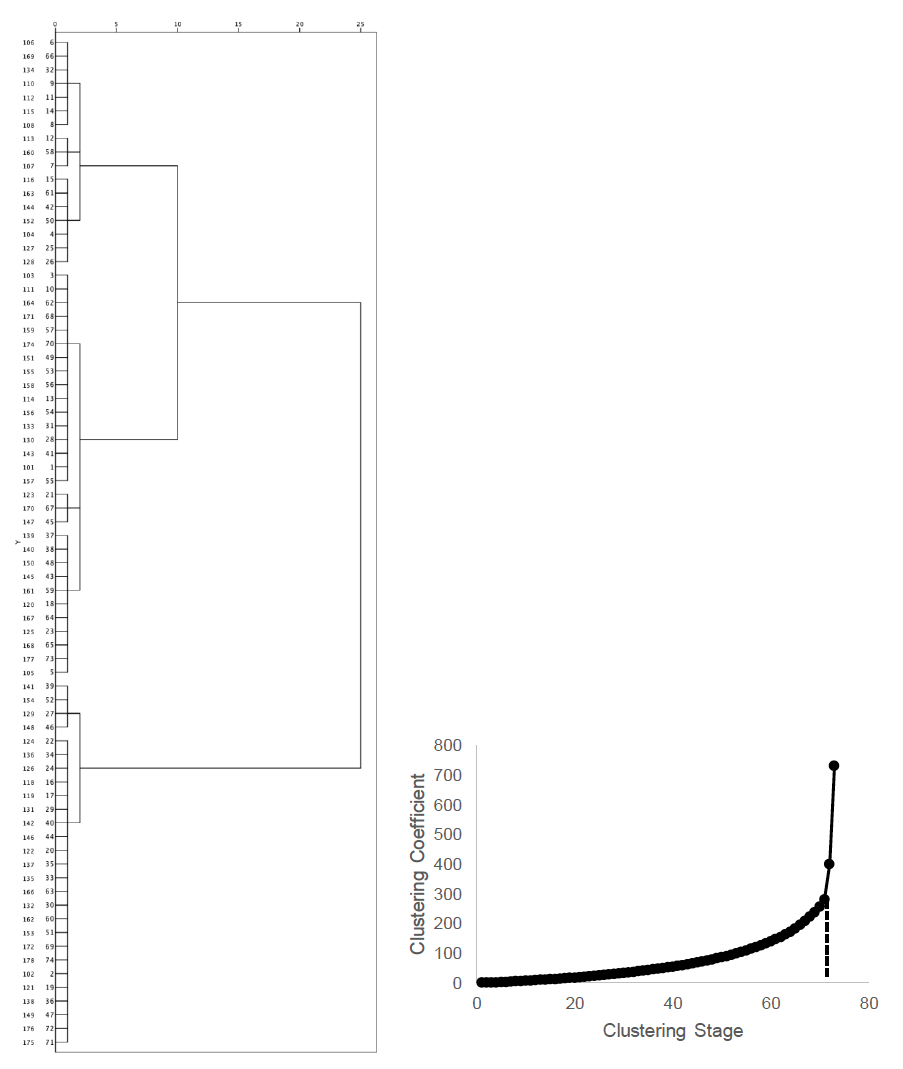

| **Supplementary Table 4**  *Initial Cluster Centers Determined from Agglomerative Hierarchical Clustering* | | | |
| --- | --- | --- | --- |
| *Z*-score | Cluster 1 | Cluster 2 | Cluster 3 |
| Expressive Vocabulary Test-3 | -.70 | .72 | -2.16 |
| CELF-5 Semantic Relationships | -1.35 | .51 | -2.36 |
| CELF-5 Word Classes | -1.03 | .72 | -2.40 |
| CELF-5 Recalling Sentences | -1.46 | -.03 | -2.52 |
| Peabody Picture Vocabulary Test-5 | -.83 | .71 | -2.51 |
| CELF-5 Understanding Spoken Paragraphs | -1.90 | -.71 | -2.55 |
| CELF-5 Sentence Assembly | -1.30 | -.01 | -2.52 |
| CELF-5 Formulated Sentences | -1.91 | -.40 | -2.64 |
| NVIQ | -.95 | .30 | -1.58 |
| Syllable Repetition Task accuracy | -.25 | .77 | -7.21 |
| *Note.* Expressive Vocabulary Test-3rd Ed. (Williams, 2018). Peabody Picture Vocabulary Test-5th Ed. (Dunn, 2018). CELF-5 = Clinical Evaluation of Language Fundamentals-5th Ed. (Wiig et al., 2013). Syllable Repetition Task (Shriberg et al., 2009). | | | |

| **Supplementary Table 5**  *Internal Clustering Criteria, with Bolded Text Indicating the Optimal Number of Clusters* | | | | | | | | |
| --- | --- | --- | --- | --- | --- | --- | --- | --- |
| Number of Clusters: | 2 | 3 | 4 | 5 | 6 | 7 | 8 | 9 |
| 1. AIC | 3111.90 | **3073.65** | 3097.68 | 3122.57 | 3149.04 | 3178.52 | 3210.38 | 3240.98 |
| 2. Average silhouette width | .529 | **.560** | .504 | .397 | .407 | .401 | .394 | .423 |
| 3. BIC | **3202.962** | 3210.252 | 3279.814 | 3350.234 | 3422.238 | 3497.255 | 3574.643 | 3650.783 |
| 4. Calinski-Harabasz | 64.053 | **66.487** | 48.788 | 39.266 | 38.267 | 32.791 | 29.138 | 26.409 |
| 5. C-index | **.182** | .177 | .151 | .138 | .135 | .133 | .127 | .127 |
| 6. Cubic clustering criterion | -2.609 | **-1.741** | -3.126 | -3.998 | -4.292 | -5.111 | -5.689 | -6.161 |
| 7. Davies-Bouldin | **.950** | 1.115 | 1.610 | 1.876 | 1.792 | 1.880 | 1.667 | 1.711 |
| 8. Dunn clustering criterion | .053 | .055 | .050 | **.073** | .071 | .073 | .073 | .073 |
| 9. Gamma statistic | **.644** | .569 | .590 | .614 | .620 | .621 | .633 | .630 |
| 10. Log determiner ratio | 1.221 | **2.333** | 2.997 | 3.654 | 4.323 | 5.040 | 5.449 | 5.882 |
| 11. Log sum of squares ratio | -.089 | **.656** | .767 | .852 | 1.064 | 1.108 | 1.159 | 1.210 |
| 12. McClain-Rao | .593 | .626 | .608 | .591 | .591 | .589 | **.581** | .581 |
| 13. PBM Index | 34.223 | **40.281** | 31.410 | 29.084 | 28.502 | 24.739 | 22.758 | 20.612 |
| 14. Point-biserial *r* | **.537** | .418 | .382 | .396 | .369 | .350 | .344 | .355 |
| 15. Pooled sum of squares within | 45846.45 | **29994.26** | 27851.07 | 26253.53 | 22518 | 21803.03 | 20969.43 | 20166.85 |
| 16. Ratakowsky-Lance Index | .439 | **.477** | .387 | .357 | .328 | .308 | .295 | .281 |
| *Note.* AIC = Akaike Information Criterion. BIC = Bayesian Information Criterion. Log sum of square ratio = “top elbow” method. PBM = Pakhira-Bandyopadhyay-Maulik. Pooled sum of squares within = “bottom up” elbow method. | | | | | | | | |

**Supplementary Figure 4.** Clustering of participants (N = 74) using a k-means three-cluster solution. Cluster 1 = circles. Cluster 2 = squares. Cluster 3 = triangles.

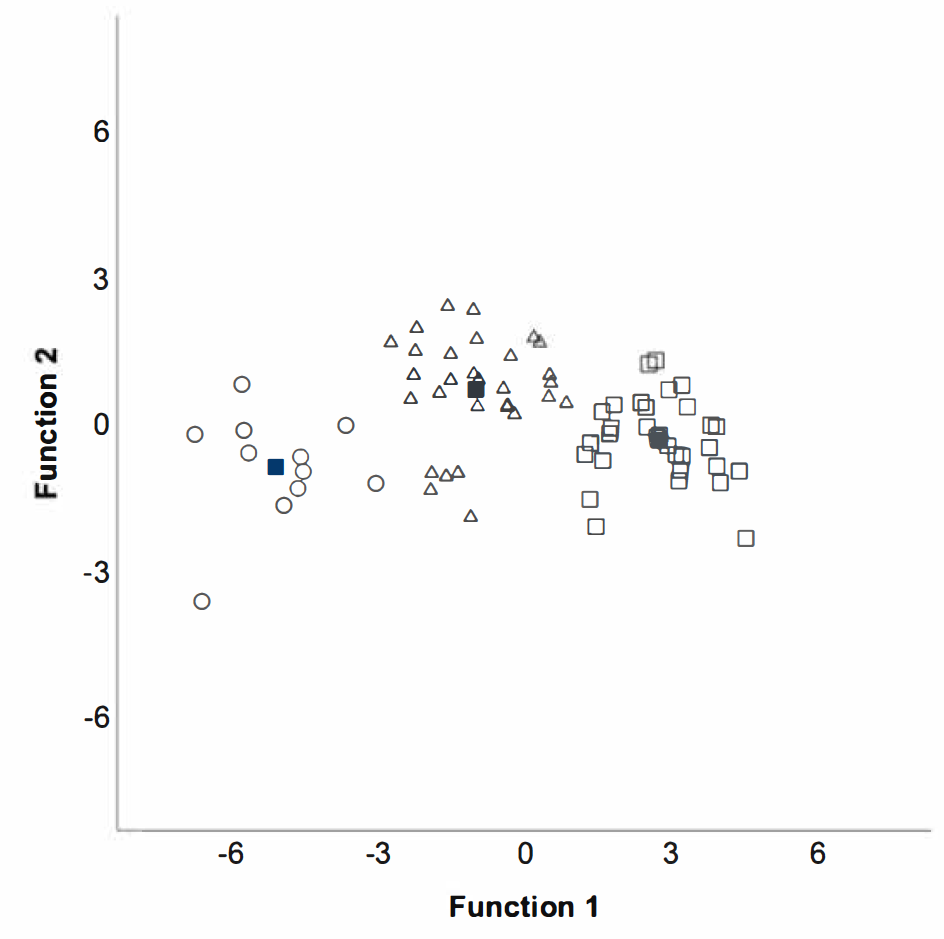
